# The impact of premature extrauterine exposure on infants’ stimulus-evoked brain activity across multiple sensory systems

**DOI:** 10.1101/2021.07.20.21260866

**Authors:** Gabriela Schmidt Mellado, Kirubin Pillay, Eleri Adams, Ana Alarcon, Foteini Andritsou, Maria M Cobo, Ria Evans Fry, Sean Fitzgibbon, Fiona Moultrie, Luke Baxter, Rebeccah Slater

**Author notes:** Corresponding author Paediatric Neuroimaging Group, Department of Paediatrics, Level 2 Children’s Hospital, John Radcliffe Hospital, University of Oxford, Oxford, United Kingdom. These authors contributed equally (Gabriela Schmidt Mellado, Kirubin Pillay). These authors contributed equally (Luke Baxter, Rebeccah Slater).

## Abstract

Prematurity can result in widespread neurodevelopmental impairment, with the impact of premature extrauterine exposure on brain function detectable in infancy. A range of neurodynamic and haemodynamic functional brain measures have previously been employed to study the neurodevelopmental impact of prematurity, with methodological and analytical heterogeneity across studies obscuring how multiple sensory systems are affected. Here, we outline a standardised template analysis approach to measure the evoked response magnitudes for visual, tactile, and noxious stimulation within individual infants (n=15) using EEG. By studying a cohort of very preterm infants longitudinally (n=10), we observe that the evoked response template magnitudes are significantly associated with age-related maturation. In a subsequent cross-sectional study, we observe significant differences in visual and tactile response template magnitudes between infants born in the very preterm and late preterm periods (n=10 and 8), age-matched at study. These findings demonstrate the significant impact of premature extrauterine exposure on brain function and suggest that prematurity can accelerate maturation of the visual and tactile sensory system in infants born very prematurely. This study highlights the value of using a standardised multi-modal evoked-activity analysis approach to assess premature neurodevelopment, and will likely complement resting-state EEG and behavioural assessments in the study of the functional impact of developmental care interventions.

**Highlights:**

- Multi-modal sensory stimulation is used to assess premature infant brain function
- A template analysis approach is outlined for multi-modal stimulus-evoked EEG activity
- Stimulus-evoked template magnitudes change with age
- Prematurity results in accelerated maturation of visual and tactile sensory systems

## 1. Introduction

Premature birth and early exposure to the extrauterine environment can result in widespread neurodevelopmental impairment, dependent on the degree of prematurity at birth and postnatal insults (Blencowe et al., 2013; Wallois et al., 2020). Relative to in-utero, the extrauterine environment increases exposure to noise, light, handling and clinically painful procedures, as well as pharmacological interventions, which all have the potential to detrimentally influence developmental trajectories (Pineda et al., 2014; Smith et al., 2011). During the third trimester, dramatic changes to both brain structure and function occur (André et al., 2010; Dubois et al., 2014; Dubois and Dehaene-Lambertz, 2015; Wallois, 2010), and in addition to brain activity being endogenously generated (Colonnese and Khazipov, 2012), neural activity becomes progressively sensory driven in anticipation of patterned environmental sensory input (Colonnese et al., 2010; Milh et al., 2007; Wess et al., 2017). Given the important role of sensory input in neurodevelopment, infants exposed to the extrauterine environment during these critical periods are vulnerable to the influence of developmentally unexpected environmental stimuli.

Prematurity can negatively impact the development of multiple sensory systems (Wallois et al., 2020). Children born prematurely can develop atypical somatosensory profiles (Adams et al., 2015; Wickremasinghe et al., 2013), decreased mechanical and thermal sensitivities related to pain exposure (Walker et al., 2009), sensorineural deafness from excessive noise and drug exposure (McMahon et al., 2012; Zimmerman and Lahav, 2013) and cortical visual impairment (Rosenberg et al., 1996; Van Braeckel and Taylor, 2013). Brain activity evoked by sensory stimulation can be recorded using electroencephalography (EEG) to assess the functional integrity of these developing sensory systems. Somatosensory (Leikos et al., 2020; Majnemer and Rosenblatt, 1996; Vanhatalo et al., 2009), noxious (Hartley et al., 2017; Slater et al., 2010b), auditory (Ribeiro and Carvallo, 2008; Taylor et al., 1996), and visual (Majnemer and Rosenblatt, 1996; Taylor et al., 1992; Whyte, 1993) evoked potentials have all previously been independently identified in infants using a variety of analytical methods.

However, to date, there is limited data assessing the impact of extrauterine exposure on the developmental trajectory of sensory evoked cortical activity across multiple stimulus modalities within the same infants. This severely limits our understanding of the impact that premature ex-utero development has across these sensory systems in terms of the relative magnitude and directionality of effect. A major challenge to assessing the impact of prematurity across multiple sensory systems is lack of statistical power due to limited sample sizes as well as limitations to the number of stimulus trials that can be applied per infant. At the extreme is assessment of noxious input to study the infant pain system, where sample sizes for infant pain studies are often limited and noxious events are often single-trial clinically-required events, such as heel lances (Courtois et al., 2016).

In this study, we assess the impact of premature extrauterine environmental exposure on the development of evoked EEG brain activity across four stimulus modalities: visual, tactile, noxious, and auditory. We first characterise these stimulus evoked responses in a cohort of healthy term infants. Due to the single-trial nature of clinically required noxious procedures, we uniformly applied a template analysis approach to each stimulus modality, which we have formerly developed and validated for the analysis of noxious-evoked brain activity in neonates (Hartley et al., 2017, 2015). We have previously shown that a template of noxious-evoked brain activity capturing the dominant evoked waveform is an analysis approach robust enough for successful application to single-trial data, sensitive to a range of biologically interesting demographic and clinical variables (Cobo et al., 2021; Green et al., 2019; Gursul et al., 2018; Hartley et al., 2021, 2016; Kasser et al., 2019; Vaart et al., 2019), and provides a sensitive primary outcome in analgesic clinical trials (Hartley et al., 2018).

Here, we systematically apply the template analysis approach to each modality of stimulus-evoked EEG data acquired from a 25-electrode array to optimise and standardise our analysis pipeline and minimise analytic heterogeneity across the stimulus modalities. In an independent cohort of infants born very prematurely, we demonstrate that the stimulus-evoked response templates are sensitive to age-related developmental maturation. Furthermore, we report that the magnitudes of visual and tactile-evoked responses significantly differ between age-matched infants born in the very and late preterm periods, demonstrating the differential impact of premature birth on the maturation of functional sensory responses.

## 2. Methods

### 2.1. Overview

#### 2.1.1. Study organisation

This study is divided into three sections. First, EEG data from a cohort of healthy term-born infants (Figure 1, Full-term) were analysed to establish the template analysis methodology that would be applied across stimulus modalities in subsequent analyses. Second, the outlined template analysis approach was applied to EEG data from an independent cohort of very preterm infants (Figure 1, Very preterm). These infants were studied longitudinally to assess the association between the stimulus-evoked EEG template magnitude and infants’ age at study. Third, the template analysis was applied to EEG data from the very preterm infants’ final recording session (Figure 1, Very preterm, >34 weeks PMA), and these stimulus-evoked EEG template magnitudes were compared to that of an independent cohort of age-matched late preterm infants (Figure 1, Late preterm). This comparison assessed the association between the stimulus-evoked EEG template magnitude and the degree of extrauterine environmental exposure.

**Figure 1:**
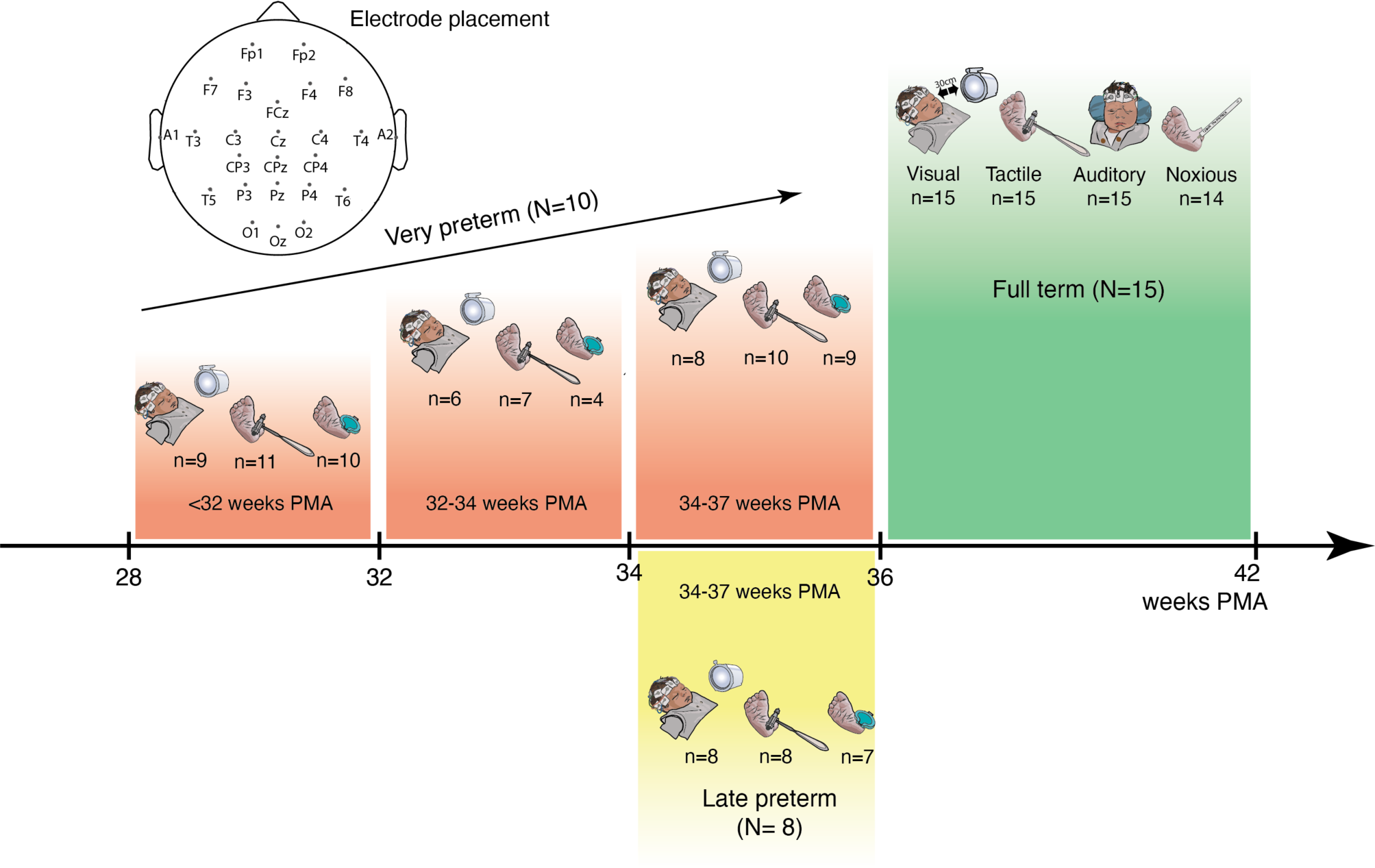
Study organisation. Three independent infant samples were assessed in this study. The healthy full-term cohort data (green) were analysed to develop the standardised template analysis pipeline (Sections 2.2 and 3.1). The very preterm cohort data (red) were analysed to assess the sensitivity of the stimulus-evoked template magnitudes to age-related maturation (Section 2.3 and 3.2). The late preterm cohort data (yellow) and final recording session of the very preterm cohort data (red, 34-37 weeks PMA) were analysed to assess the sensitivity of the stimulus-evoked template magnitudes to premature extrauterine exposure (Section 2.4 and 3.3). N = number of subjects; n = number of trial average recordings per stimulus included in the analysis; PMA = postmenstrual age (gestational age + postnatal age). Top: electrode arrangement.

#### 2.1.2. Participant recruitment

Infants born between February 2018 to February 2020 from the Maternity Unit and Special Care Baby Unit at the John Radcliffe Hospital, Oxford, UK, were enrolled in this study after informed, written consent was obtained from the parents. Ethical approval was obtained from an NHS Research Ethics Committee (National Research Ethics Service, REC reference: 12/SC/0447), and research was conducted in accordance with Good Clinical Practice guidelines and the Declaration of Helsinki. Each participant was eligible to take part in the study if they had no major neurological malformations or conditions such as intraventricular haemorrhage (IVH) grade 2 or higher, and no history of maternal substance abuse. At the time of each study, infants were clinically stable, did not require invasive ventilation, and had received no sedatives or analgesics within the prior 24 hours. Each cohort is described below, and the full demographic information is detailed in Table 1.

**Table 1:**
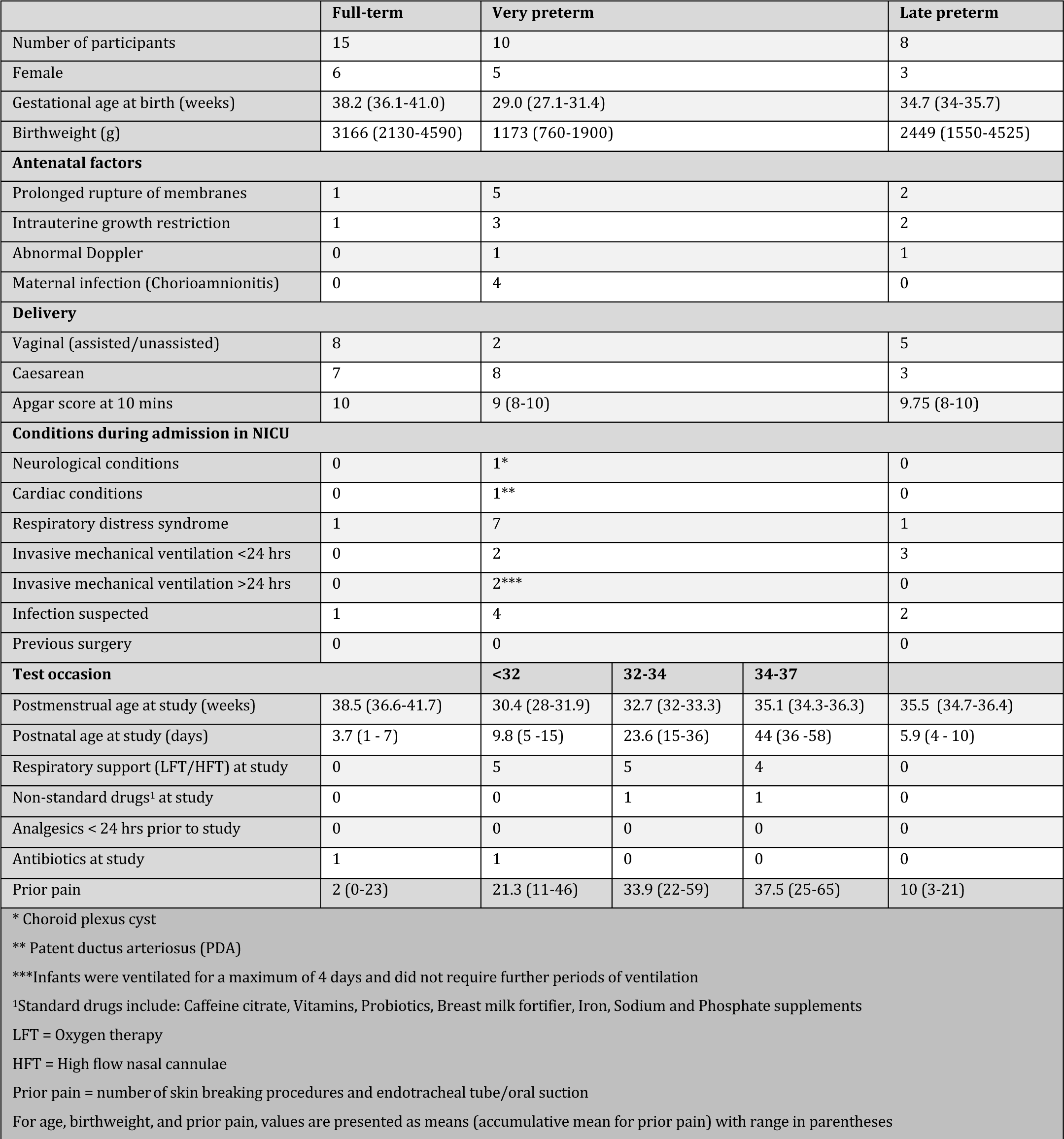
Infant demographics for the three independent cohorts used in this study.

#### 2.1.3. EEG setup

All EEG data were recorded with a sampling frequency of 2 kHz using a 25-recording electrode arrangement and CURRY scan7 neuroimaging software suite. Channels included Fp1, Fp2, F3, F4, F7, F8, FCz, T3, T4, C3, C4, Cz, CP3, CP4, CPz, T5, T6, P3, P4, Pz, O1, O2, Oz, A1 and A2 electrodes, with Fz as reference and FPz as ground. Electrodes were placed according to the 10-20 International Electrode Measuring System, and recordings acquired by a trained clinical neurophysiologist (GSM).

#### 2.1.4. Sensory stimulation

After allowing infants to settle and verifying EEG recording quality, each stimulus was applied in a consecutive train of 20-30 trials, allowing at least 10 seconds between trials. Visual, tactile, auditory and noxious stimuli were each applied in a randomised order apart from the clinical procedure i.e. the heel lance, which was performed first or last depending on clinical requirement. Due to technical issues, it was not always possible to perform all stimuli on every individual, and the final numbers of stimuli applied are summarized in Figure 1.

The visual stimulus was a flash of light presented using a Lifelines Photic Stimulator (intensity level 4, approximately 514 lumens), which was manually held 20-30 cm from the infant’s eyes. Light flashes automatically triggered event marking of the EEG recording. The tactile stimulus was a modified tendon hammer with a 154 mm diameter contact surface applied gently to the infant’s heel. The modified tendon hammer includes a built-in force transducer, which triggered an event mark on the EEG monitor (Worley et al., 2012).

Experimental or clinical noxious stimuli were applied depending on the cohort of infants and clinical procedures required. The two preterm cohorts (Figure 1, Very preterm and Late preterm) had a clinical noxious stimulus in the form of a heel lance (BD Quikheel Preemie Lancet – Becton, Dickinson and Company) as they clinically required blood sampling at the time of study. The heel lance pierces the skin at a depth of 0.85 mm, and no lance was applied solely for the purpose of this study. The heel lance was only applied once during each recording resulting in a single trial per recording. In the healthy full-term cohort (Figure 1, Full-term), no infants required a clinical noxious stimulus, so a mild 128 mN experimental sharp-touch stimulus (PinPrick Stimulator, MRC Systems) was applied to the heel of the foot instead (20-30 trials). The pinprick stimulus produces EEG responses of lower amplitude than those of clinical procedures, such as the heel lance, and does not cause behavioural distress (Hartley et al., 2015) but activates nociceptors in the periphery (Magerl et al., 2001) and elicits noxious-evoked brain activity (Baxter et al., 2021; Iannetti et al., 2013; Slater et al., 2010b). Both the experimental pinprick and clinical heel lance stimuli had built-in interfaces to trigger an EEG event mark.

The auditory stimulus was a fixed tone at 1,000 Hz frequency, delivered using a pair of speakers (n=5 late preterm cohort, n=3 very preterm cohort) or headphones placed firmly on the infant’s ears (n=3 late preterm cohort, n=5 very preterm cohort). Headphones were used for all the infants in the full-term cohort. The speakers (X-mini MAX II Portable Speakers, Xmi) were placed 15 cm from the ears and delivered the tone with 100 ms duration at a volume of 80 dB. The headphones delivered the tone with 4 ms duration and a volume of 70 dB nHL using the Surpass Ltd (EMS Biomed) auditory system.

#### 2.1.5. EEG data preprocessing

Each individual stimulus trial was first epoched in the period ±5 s either side of the stimulus and temporally filtered using 0.5-70 Hz bandpass filter and a 50 Hz notch filter. Data were average referenced to improve the spatial distribution of the activity, and to ensure that subsequent analyses were not being driven by frontal activity from the reference electrode (i.e. Fz). Electrodes in positions more likely to be affected by biological artefacts such as eye blinks (Fp1 and Fp2), muscle and ECG artefact (A1, A2), and those which were not part of the standard 10-20 electrode positions (FCz, CP3, CPz, CP4) were excluded when determining the average.

All trials were then visually assessed for quality by a trained clinical neurophysiologist (GSM). Trials were rejected only if the pre-stimulus baseline or the post-stimulus activity contained any detrimental artefacts across channels due to subject motion or electrode drop-off, or if the pre-stimulus baseline was unsettled. Of 2,954 trials, 1,874 (visual=761, tactile=903, pinprick=210) were retained. An average of 24 trials (range 14-30 trials) were performed per stimulus and a mean of 9 (range 2-20 trials) rejected from a single recording. A single trial heel lance was recorded in all the infants from the very preterm (28 recordings) and late preterm (8 recordings) cohorts. In total, 6 heel lance trials out of 36 were rejected due to artefact.

The average of the retained trials (with the exception of the single-trial heel lance stimulus) was used to produce a single stimulus-evoked response per infant recording and stimulus. Spherical interpolation was performed on the trial-averages to correct any channels that contained remaining artefacts, largely due to involuntary blinking when the visual stimulus was applied. In each case, a maximum of four channels (not adjacent to each other) were interpolated per recording and a Laplacian arrangement (whereby each channel was considered adjacent to up to four neighbouring channels according to position on the head) was used to determine which electrodes were considered adjacent to one another. All further analyses were done using the trial average per infant and stimulus, which was additionally baseline-corrected using the pre-stimulus mean in the 500 ms immediately before application of the stimulus.

### 2.2. Developing a standardised template analysis approach

#### 2.2.1. Participant details

A standardised template analysis approach was developed using the full-term infant dataset (Figure 1, Full-term; n=15; mean GA at birth = 38.2 weeks; mean PMA at study = 38.5 weeks). Each individual was studied on a single occasion within the first 15 days from birth. See Table 1 for full demographic details.

For assessing noxious stimulus-evoked brain activity, a noxious-evoked response template was derived using the full-term infant dataset collected during the present study and used throughout, rather than using the previously established noxious-evoked response template (Hartley et al., 2017). This ensured the template analysis approach used in this study was applied consistently for all stimulus modalities, due to some methodological differences between the current study and the previous study’s approach to deriving a noxious-evoked template e.g. the current full-term dataset was collected using higher electrode density and greater number of noxious stimulation trials, and was analysed using an average reference rather than referenced to Fz.

#### 2.2.2. Electrode selection using functional neuroanatomical considerations and raw amplitude analysis

To evaluate the brain region or electrode position where maximal amplitude change occurs after application of a stimulus, the evolution of the spatial distribution of amplitude changes immediately after the stimulus was visually assessed. This was done using a grand average of the trial averages per infant for each stimulus. Using a priori functional neuroanatomical considerations, large evoked responses were expected around Oz for visual stimulation, Cz for tactile stimulation of the foot, and T3 and T4 for auditory stimulation (Vanderah and Gould, 2020); and from previous publications on EEG activity evoked by the specific noxious stimuli used in the present study, large evoked responses were expected around Cz (Hartley et al., 2017; Slater et al., 2010b). The pre-processed EEG amplitude changes were visually evaluated to assess these prior expectations and the qualitative biophysical interpretability of the recorded data. If the evoked raw amplitude data were consistent with these prior expectations, these functional neuroanatomical considerations and previous publications formed the basis for electrode selection of a single electrode.

#### 2.2.3. Time window selection using temporal cluster analysis

Having selected the electrode where maximum response amplitude occurred for each stimulus modality, we identified the time window during which the evoked deflections were significantly greater than the pre-stimulus background activity. Using the trial- and subject-averaged timeseries, a time window within the first 800 ms post-stimulation containing a single positive or negative deflection was identified using non-parametric temporal cluster analysis (Maris and Oostenveld, 2007). Prior to the temporal cluster analysis, all trial-averages were additionally baseline-normalised (dividing by the pre-stimulus standard deviation 500 ms immediately before the stimulus) in order to minimize discrepancies in the trial-averages due to age-related variance in signal amplitude.

For the temporal cluster analysis, each timepoint in the post-stimulus period was compared to the pre-stimulus period of resting activity across all infants’ trial-averaged EEG based on a paired t-test at the specific channel of interest. All timepoints exceeding the t-statistic threshold of 97.5% were considered significant and time-adjacent values were grouped into clusters. The cluster masses (sum of the t-statistics within each cluster) were compared against a permutation distribution of cluster masses derived after repeating this procedure with the post-stimulus and pre-stimulus labels randomly re-assigned each time for 1,000 iterations. Remaining clusters whose mass exceeded the permutation distribution in a two-tailed test (p<0.025) were considered significant. The significant post-stimulation time window was defined as the time range of the first and last timepoint in the significant cluster.

#### 2.2.4. Waveform selection using PCA and cross-correlation analysis

We used principal component analysis (PCA) to derive the waveforms that characterised the dominant post-stimulus activity for each stimulus within the stimulus-specific time windows identified using the temporal cluster analysis (section 2.2.3.). For each stimulus modality, the EEG periods across all channels within the specified windows were concatenated together before PCA, and the first three components were considered. We used singular value decomposition (SVD) to project each component onto the data and scale each component waveform for best fit. The scaling factor is the component magnitude. When fitting the component waveforms using SVD, latency differences between individuals were corrected for using Woody filtering (Woody, 1967), which jitters the signal relative to the component to achieve maximum cross-correlation. A maximum jitter of ±50 ms was selected for visual and noxious stimuli and ± 100 ms for the tactile stimulus (no significant time windows were identified for auditory stimuli). For each component, the cross-correlation between the fitted component waveform and the evoked data was calculated, and the component waveform with the largest cross-correlation within a stimulus modality was selected as the stimulus-evoked response template.

We generated surface heat maps to qualitatively assess the spatial topography of each template’s magnitude by fitting the templates to all electrodes. The Woody filtering jitter estimated at each stimulus modalities dominant electrode (identified in section 2.2.2.) was used across all channels. An additional background correction was applied independently for each channel to reduce the effects on the template’s magnitudes brought about by changes in the underlying background activity of the signal (such as variations in sleep state) between stimuli and individuals. After estimating the template magnitude in the post-stimulus period, the same procedure was applied to successive windows of stable pre-stimulus baseline activity (with length matching the template duration) across the 1 s period prior to the stimulus. The median of these values resulted in a robust pre-stimulus template magnitude as a surrogate for the background response magnitude and was then subtracted from the stimulus-evoked response template magnitude to obtain a corrected value.

#### 2.2.5. Applying the standardised template analysis approach to novel data

Each stimulus-evoked response template was rescaled such that the average template magnitude for the full term cohort used to develop the template was a unit vector i.e. a template magnitude equal to one is the expected value for full term healthy infants. The stimulus-evoked response templates selected in section 2.2.4. were fit to new stimulus-evoked EEG data using the electrode selected in section 2.2.2. and the time window selected in section 2.2.3. The fit was performed using SVD to efficiently perform a least squares optimisation, and maximum Woody filtering jitters of ±50 ms for visual and noxious stimuli and ±100 ms for tactile stimuli. The template magnitudes were corrected using the median pre-stimulus template magnitude, as described in section 2.2.4.

### 2.3. Detecting age-related developmental changes in stimulus-evoked brain activity using the standardised template analysis approach

#### 2.3.1. Participant details

Assessment of the correlation between stimulus-evoked template magnitudes and infant age was performed using the longitudinally studied very preterm dataset (Figure 1, Very preterm; n=10; mean GA at birth = 29 weeks). Infants were studied on average three times before discharge from hospital (range 2-4 test occasions). The recordings were later separated into three age groups: <32 weeks (mean PMA at study = 30.4 weeks), 32-34 weeks (mean PMA at study = 32.7 weeks), 34-37 weeks (mean PMA at study = 35.1 weeks). See Table 1 for full demographic details.

The infants in this cohort were remarkably healthy preterm infants given their young age at birth. A single infant had a minor neuroimaging finding on their ultrasound scan (choroid plexus cyst) of no pathological significance, and that same infant also had a minor cardiac condition. Four infants received mechanical ventilation during their first week of extrauterine life for a short period of time, but not at the time of study. The incidence of chronic lung disease could not be assessed when the last test occasion was performed (infants were too young). One infant was severely IUGR (intra-uterine growth restricted) at the first test occasion and had a dysmature background EEG that improved in the subsequent test occasions. The initial abnormal recording was therefore excluded from the analysis. The first test occasion was performed within 15 days of birth and repeated once every 2 weeks thereafter until near discharge from hospital, resulting in 28 recordings. Each study was arranged to coincide with a clinically required heel lance, and no treatments (i.e. phototherapy, humidity protocols) or routine cares were disrupted by the studies.

#### 2.3.2. Analysis

The template analysis approach was applied to all infants’ stimulus-evoked EEG data, as outlined in section 2.2.5. We used a linear random effects regression model to correlate the stimulus-evoked template magnitudes with infant PMA. Random intercepts were introduced to group repeated recordings from the same patient. GA and the number of trials in each recording’s trial average were also separately correlated with the PMA to assess if they were potential confound variables, and no significant correlations were observed for each stimulus.

### 2.4. Detecting the impact of premature extrauterine development on stimulus-evoked brain activity using the standardised template analysis approach

#### 2.4.1. Participant details

The impact of premature extrauterine exposure on stimulus-evoked template magnitudes was assessed by comparing responses between very preterm and late preterm infants, both groups matched for age at time of study. The very preterm group was a cohort of infants born <32 weeks GA and studied at 34-37 weeks PMA (n=10; mean GA at birth = 29 weeks; mean PMA at study = 35.1 weeks; mean postnatal age at study = 44 days). This sample is the same group analysed in the longitudinal study above (see section 2.3.1 and Figure 1, Very preterm, >34 weeks). See Table 1 for full demographic details.

The late preterm group was a cohort of infants born at 34-36 weeks GA and studied at 34-37 weeks PMA (n=8; mean GA at birth = 34.7 weeks; mean PMA at study = 35.5 weeks; mean postnatal age at study = 5.9 days). This sample is an independent sample not used in any previous analyses. Infants had no known neurological or cardiac conditions at the time of study and mechanical support, when given, was not for a prolonged period of time. See Table 1 for full demographic details.

#### 2.4.2. Analysis

The template analysis approach was applied to all infants’ stimulus-evoked EEG data, as outlined in section 2.2.5. We compared the stimulus-evoked template magnitudes between the PMA-matched very preterm and late preterm infants using a linear random effects regression model. Random intercepts were applied to account for repeated measures in the case of multiple recordings from individuals within a group. Group differences in PMA and the number of trials in each recording’s trial average were also separately assessed as potential confound variables, and no significant correlations were observed for each stimulus.

## 3. Results

### 3.1. A standardised template analysis approach for extracting EEG stimulus-evoked response features across stimulus modalities in infants

#### 3.1.1. Electrode selection using raw amplitude analysis

In a cohort of full-term healthy infants (n=15; mean GA at birth = 38.2 weeks, mean PMA at study = 38.5 weeks), we applied 20-30 trials of visual, tactile, noxious, and auditory stimuli and recorded the stimulus-evoked brain activity using EEG with 25 recording electrodes. To identify the electrodes recording the strongest stimulus-evoked responses, we generated trial- and subject-averaged activity per electrode (Figure 2). For each modality, large response amplitudes were observed over cortical regions functionally-relevant to the stimulus type. A large evoked response was localised around the midline occipital electrode, Oz, after visual stimulation (Figure 2 row 1); around the midline central electrode, Cz, after both tactile and noxious stimulation (Figure 2 row 2-3); and bilaterally around temporal electrodes T3 and T4 after auditory stimulation, with larger amplitudes over the left temporal area, T3 (Figure 2 row 4).

**Figure 2:**
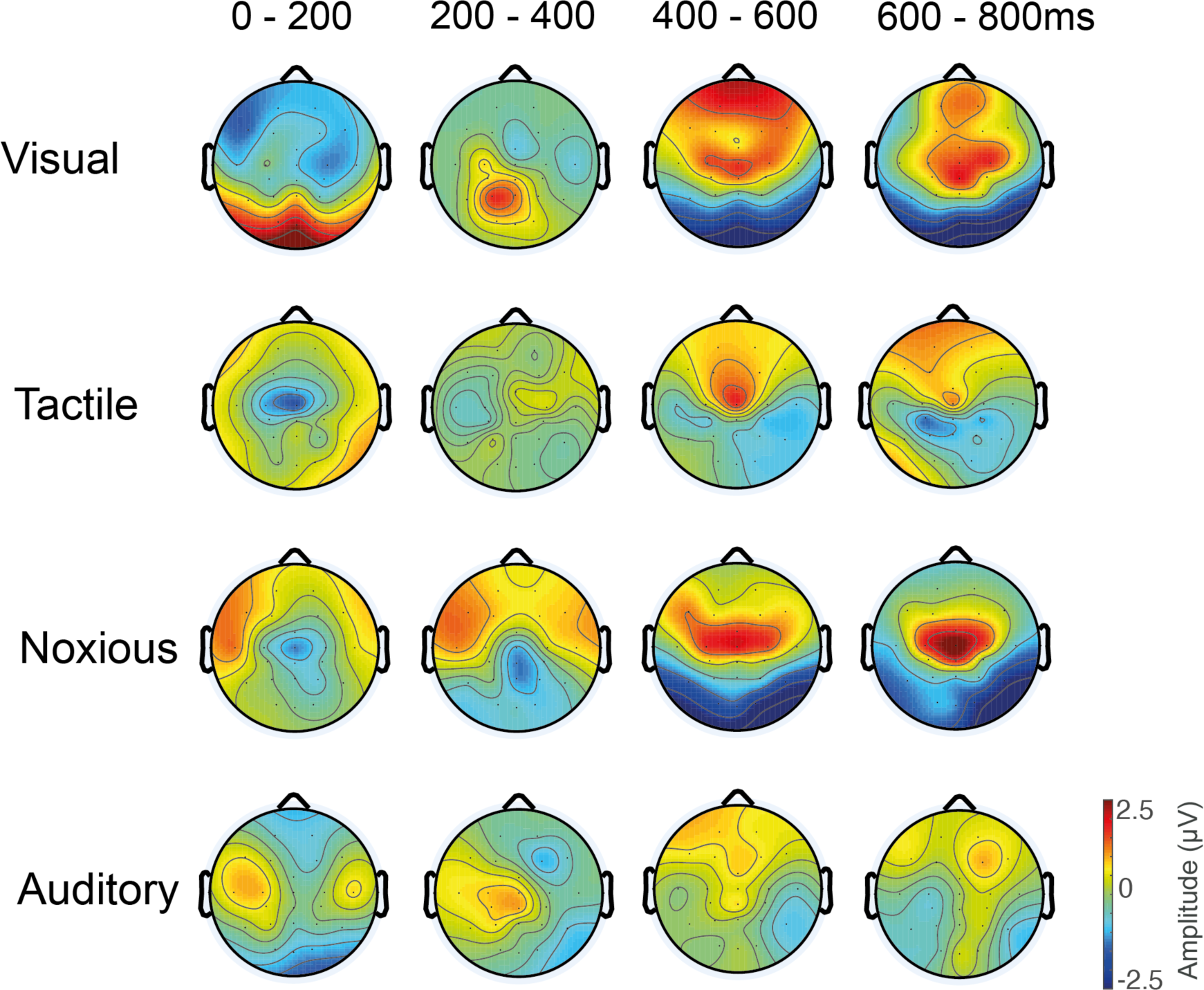
Electrode selection from maximum amplitude responses and spatial evolution of the stimulus-evoked activity over the first 800 ms post-stimulus. Changes in the raw spatial activity is presented in 200 ms intervals for each of the stimulus modalities. Visual stimulation evoked large response amplitudes at electrode Oz, both tactile and noxious stimulation evoked large response amplitudes at electrode Cz, and auditory stimulation evoked mild response amplitudes at electrode T3.

We assessed the stimulus-evoked EEG response amplitudes in successive 200 ms intervals to demonstrate the spatial evolution of the raw amplitude evoked activity immediately post-stimulus. The visual-evoked activity at electrode Oz featured a positive deflection within the first 200 ms, which evolved into a negative deflection by 400 ms. The tactile-evoked activity at electrode Cz featured a negative deflection within the first 200 ms, which evolved into a smaller amplitude positive deflection by 400 ms. The noxious-evoked activity at electrode Cz featured a modest negative deflection within the first 200 ms, which evolved into a larger positive deflection by 400 ms. And the auditory-evoked activity at electrode T3 featured a modest positive deflection within the first 200 ms.

#### 3.1.2. Time window selection using temporal cluster analysis

Having identified the maximum response amplitude electrode for each stimulus modality, we next identified the time window during which these evoked deflections were significantly greater than pre-stimulus background activity. Using a cluster analysis on the trial- and subject-averaged timeseries, a time window within the first 800 ms post-stimulation was identified, containing a single positive or negative deflection, for the visual, tactile, and noxious stimulus modalities (Figure 3). No statistically significant time window was identified for the auditory-evoked response.

**Figure 3:**
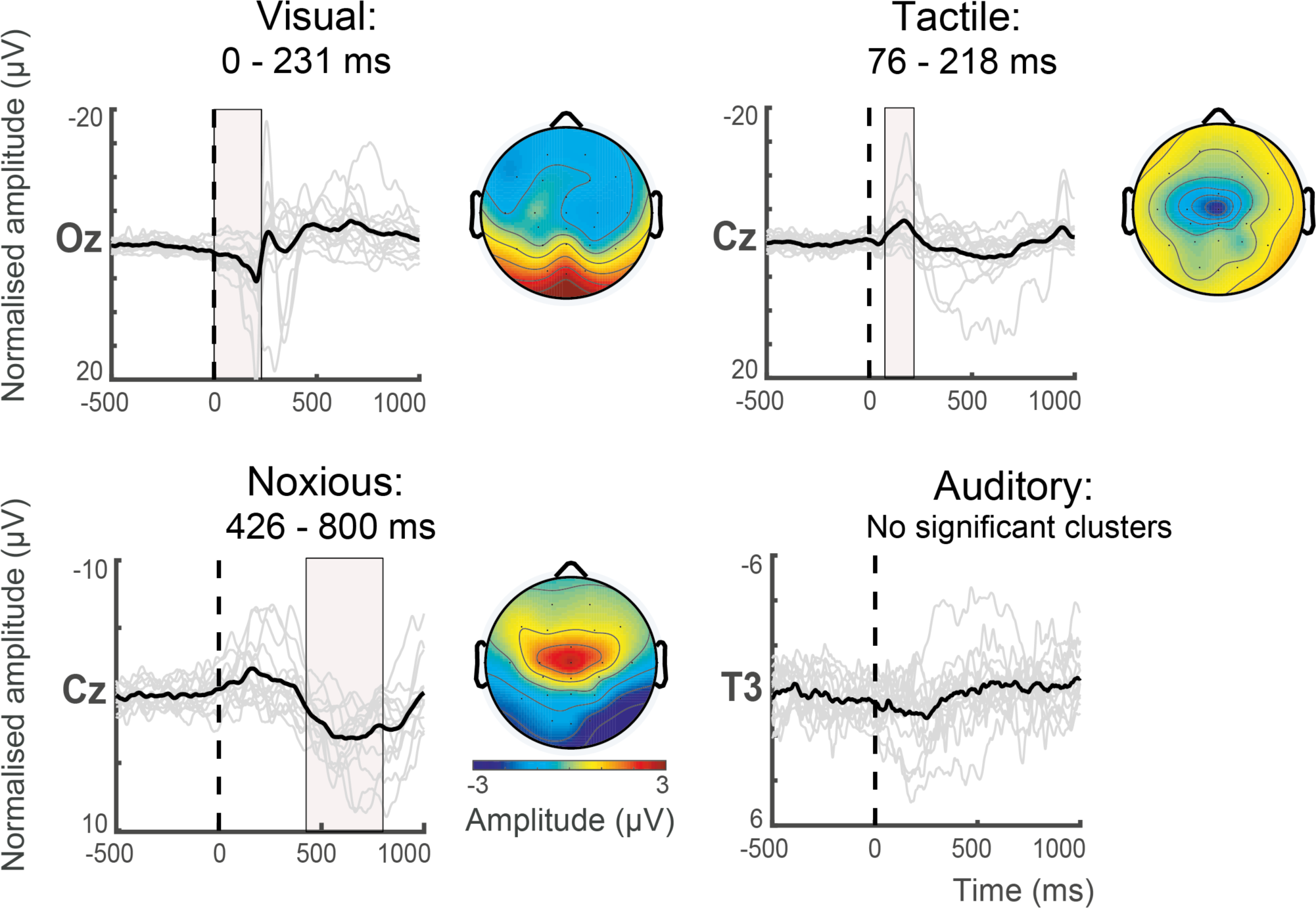
Time window selection of significant deflections in average stimulus-evoked responses at the spatial location of maximum response amplitude. The trial- and subject-averaged activity is displayed from 500 ms pre-stimulus to 1000 ms post-stimulus. Vertical dotted lines denote the point of stimulus application, solid grey lines are the individual infant (trial-averaged) responses, and the subject-averaged response is superimposed in black. The pink shaded regions denote the statistically significant temporal window spanning a positive or negative deflection in the evoked responses, identified using temporal cluster analysis. Visual–evoked responses at Oz have a significant cluster in the window 0-231ms (top left), tactile-evoked responses at Cz have a significant cluster in the window 76-218ms (top right), and noxious-evoked responses at Cz have a significant cluster in the window 426-800ms (bottom left). No significant cluster was identified for the auditory-evoked responses clusters at T3 (bottom right). For each stimulus modality, heat maps display the corresponding spatial distribution of the average EEG activity for each period showing a significant cluster. No heatmap is shown for the auditory stimulus as no significant clusters were identified.

For visual-evoked activity at electrode Oz, the 0-231 ms time window contained a significant positive deflection (p=0.001). For tactile-evoked activity at electrode Cz, the 76-218 ms time window contained a significant negative deflection (p=0.015). And for noxious-evoked activity at electrode Cz, the 426-800 ms time window contained a significant positive deflection (p=0.001).

#### 3.1.3. Waveform selection using PCA and cross-correlation analysis

Using the temporal windows containing statistically significant stimulus-evoked responses, we extracted waveforms explaining high proportions of signal variance using principal component analysis (PCA). The dominant waveform morphology per stimulus was then identified as the component with the greatest cross-correlation with the evoked activity. For all three stimulus modalities, the second component (PC2) had the greatest cross-correlation with the average EEG waveform (Figure 4). For visual-evoked activity, the first three components explained 95.7% of the variance with scaled cross-correlation values of 19.6, 21.5, and 16.8. For tactile-evoked activity, the first three components explained 87.5% of the variance with scaled cross-correlation values of 2.94, 2.99, and 1.67. And for noxious-evoked activity, the first three components explained 88.8% of the variance with scaled cross-correlation values of 43.1, 59.8, and 42.9.

**Figure 4:**
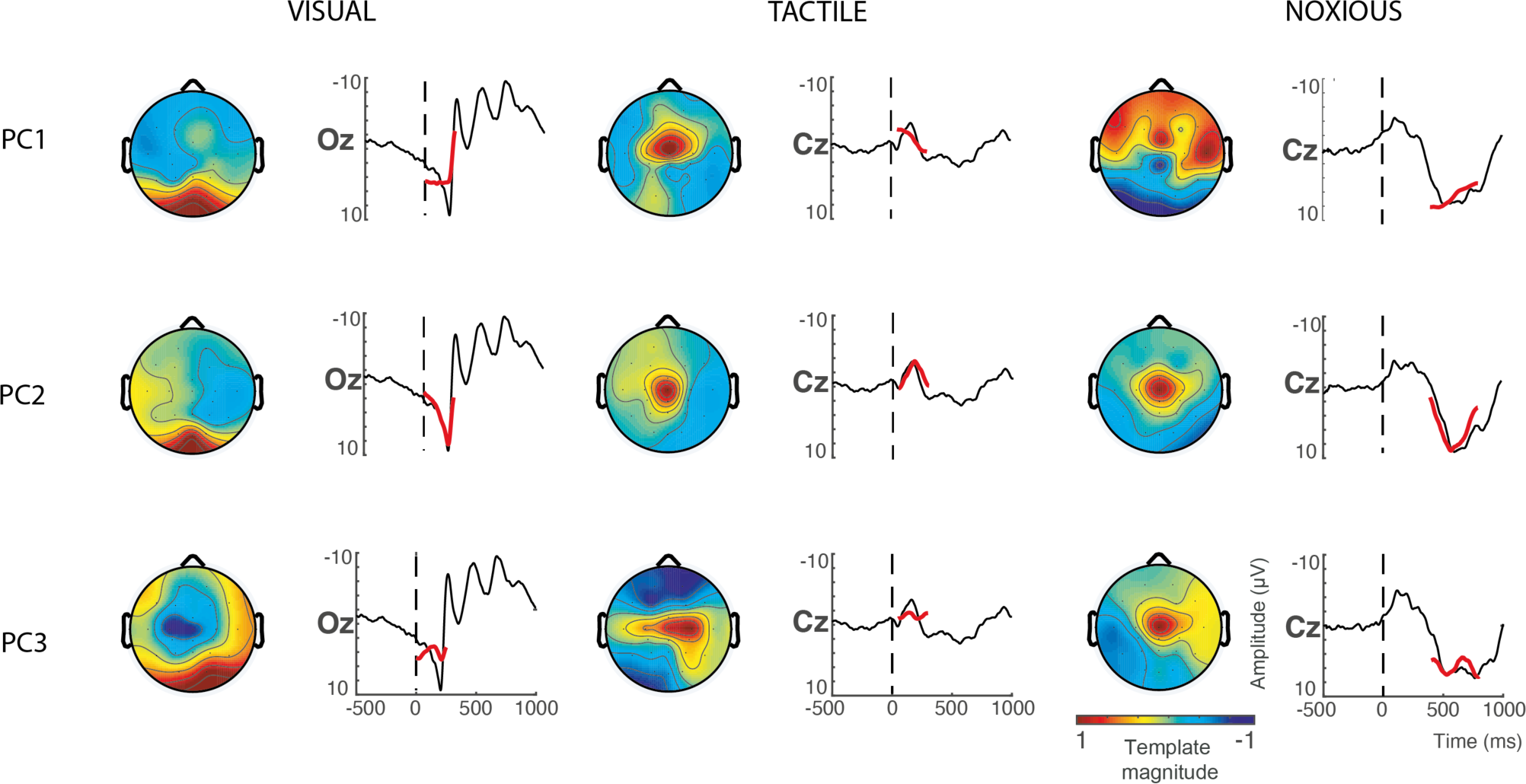
Waveform selection using PCA and cross-correlation analysis. For each stimulus modality (columns), the first three principal components PC1-3 (rows) are re-projected onto the average EEG timeseries to assess goodness-of-fit. The component’s waveforms are in red, and the average EEG timeseries are in black. Vertical dotted lines denote the point of stimulus application. For each stimulus modality, PC2 was selected as the stimulus-evoked template as it has the best fit to the EEG data (quantified using cross correlations –see main text), demonstrating clear correspondence to a positive or negative deflection.

To assess the biophysical interpretability of the selected PC2 waveforms, the three components for each stimulus modality were re-projected onto the data (Figure 4). To incorporate the full temporal morphology of the waveforms and improve generalisability to novel datasets, the temporal window in which the components were fit were widened: the visual temporal window was widened from 0-231 ms to 0-250 ms; the tactile temporal window was widened from 76-218 ms to 50-300 ms; and the noxious temporal window was widened from 426-800 ms to 400-800 ms. Goodness of fit between the component’s waveforms and the average EEG activity timeseries were assessed visually, and consistent with the quantitative cross-correlation results, PC2 qualitatively captured the dominant evoked deflection with highest fidelity. The visual template fit a positive deflection with a mean template peak latency of 205 ms, the noxious template fit a positive deflection with a mean template peak latency of 576 ms, and the tactile template fit a negative deflection with a mean template peak latency of 181 ms.

#### 3.1.4. Summary of standardised template analysis pipeline for EEG stimulus-evoked feature extraction

The three stages for feature extraction (sections 3.1.1 to 3.1.3) were applied consistently across stimulus modalities and identified biophysically sensible template waveforms for visual-, tactile-, and noxious-evoked EEG activity (Figure 5). These template vectors can be projected onto novel independent data and scaled for best fit using the spatial and temporal constraints identified in this analysis pipeline outline: visual template applied to electrode Oz within the 0-250 ms post-stimulation temporal window; tactile template applied to electrode Cz within the 50-300 ms post-stimulation temporal window; and noxious template applied to electrode Cz within the 400-800 ms post-stimulation temporal window.

**Figure 5:**
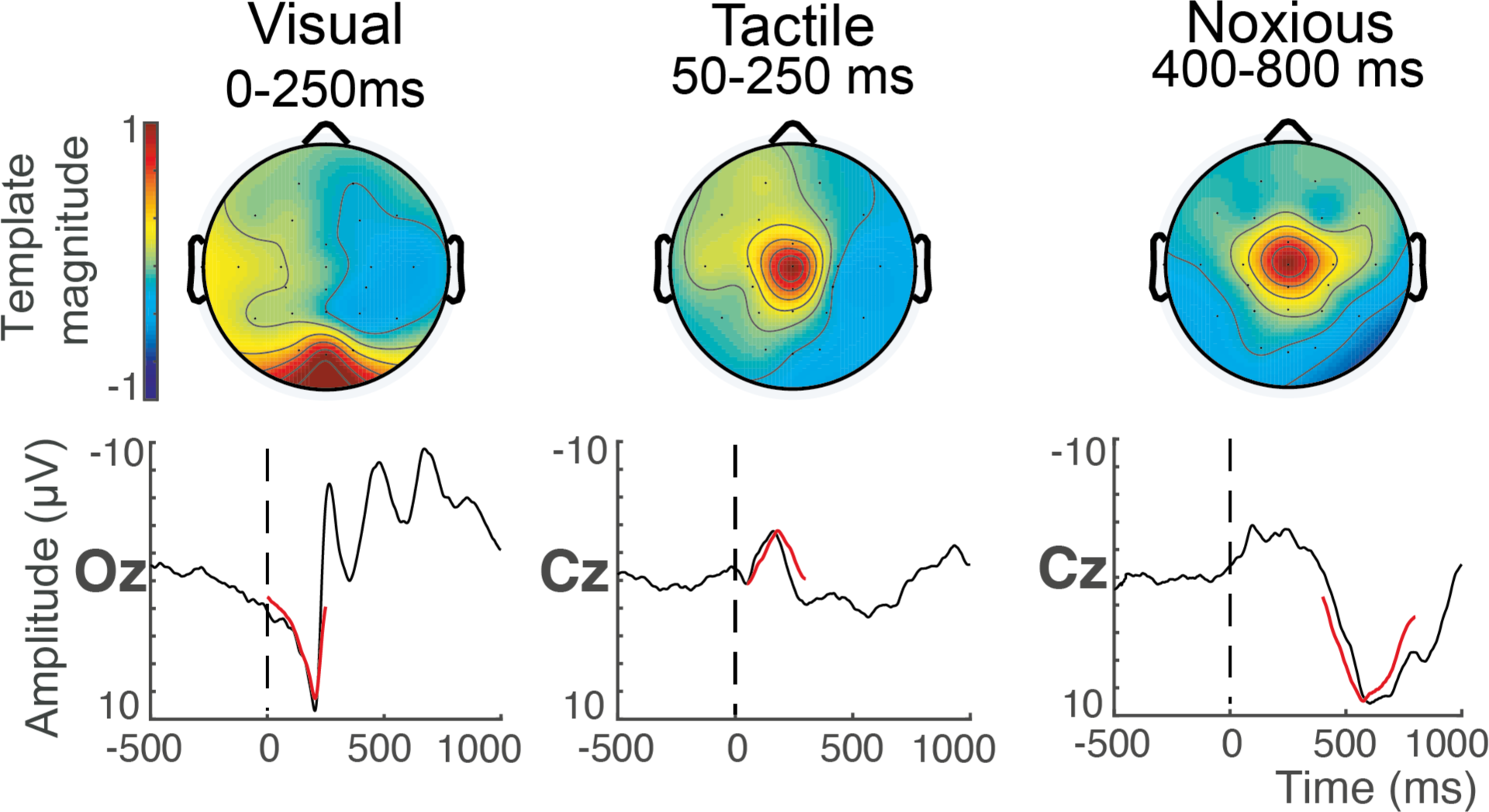
Summary of EEG template analysis pipeline results. The heat maps display the spatial localisation of average stimulus-evoked response magnitudes after re-projection onto the data across stimulus modalities. The timeseries shows the derived templates (in red) projected onto the average EEG response (black) at the maximum amplitude electrode within the stimulus-specific time windows. Vertical dotted lines show the point of stimulus application. These data were used to derive the templates, so overfitting will occur. They are displayed for qualitative summary purposes.

This scaling factor (vector magnitude) captures the response magnitude of the positive or negative deflection within the defined time windows (with a Woody filtering jitter). While these templates do not capture the entire evoked response waveform, this analysis approach assesses a prominent aspect of the response that exhibits high statistical power, a valuable property in newborn developmental EEG studies with limited sample sizes and limited stimulus trials e.g. single-trial clinically required painful procedures. In addition to magnitude, the template analysis approach can be used to extract non-magnitude features for the relevant deflection such as template latency and goodness-of-fit effects. When applying the templates to independent datasets in the following sections, we focus on magnitude effects due to existing EEG literature highlighting the biological and clinical value of stimulus-evoked response size across stimulus modalities (Hartley et al., 2016; Nevalainen et al., 2008; Whyte et al., 1987).

### 3.2. Age-related developmental changes in stimulus-evoked brain activity are detectable using template magnitudes

Using the spatial and temporal constraints outlined above (Figure 5), we applied the visual, tactile, and noxious templates to stimulus-evoked EEG data in an independent longitudinally studied cohort of very preterm infants (n=10, mean GA at birth = 29 weeks). Infant recordings were separated into three groups according to PMA: <32 weeks (n=11, mean PMA at study = 30.4 weeks), 32-34 weeks (n=7, mean PMA at study = 32.7 weeks), 34-37 weeks (n=10, mean PMA at study = 35.1 weeks). For each stimulus modality, the evoked response template magnitudes were significantly correlated with PMA with variable correlation polarities (Figure 6A). The visual-evoked response magnitudes were positively correlated with age (R^2^=0.234, p-value=0.015), as were the noxious-evoked response magnitudes (R^2^ =0.498, p-value =0.009). Conversely, the tactile-evoked response magnitudes were negatively correlated with age (R^2^=0.168, p-value =0.031).

**Figure 6:**
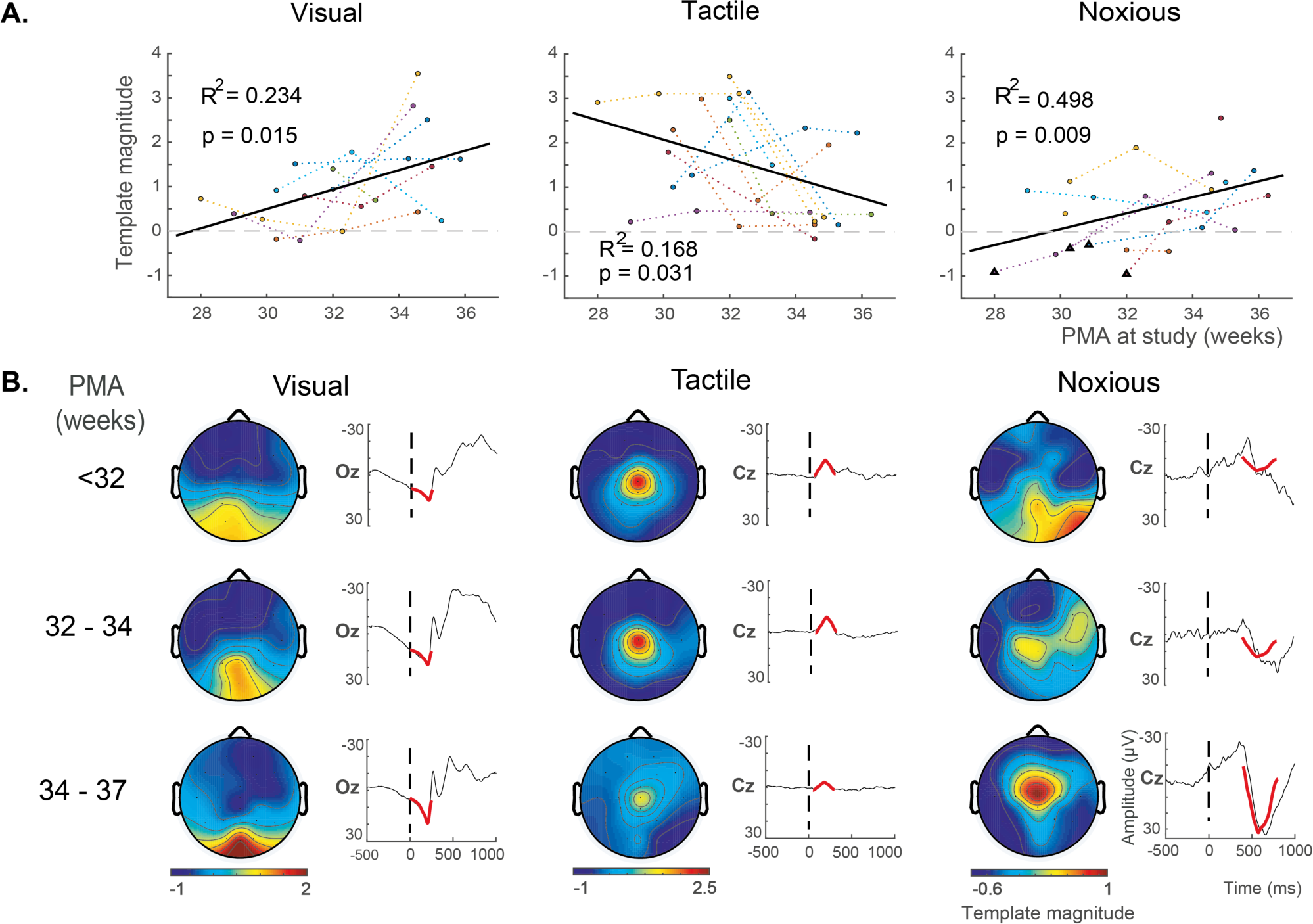
Longitudinal assessment of age-related developmental changes in stimulus-evoked response template magnitudes. (A) Correlations between template magnitudes and PMA, assessed at the dominant electrode (Oz for visual, Cz for tactile and noxious) using the template analysis approach developed in an independent cohort. Correlations were assessed using linear random effects models. Dotted lines indicate each infant’s trajectory plotted in a different colour, with the overall regression line in solid black. The dashed horizontal line denotes the point of zero magnitude. R^2^ is the regression coefficient of variation (proportion of variance explained), and p is the associated p-value. The triangle markers below the horizontal zero line of the noxious response plot indicate instances of stimulus-evoked delta brush responses. (B) Heat maps display the developmentally changing spatial patterns of the stimulus-evoked deflections, for qualitative assessment across age groups. The timeseries display the templates (in red) superimposed onto the average EEG response (black) 500 ms pre-stimulus and 1000 ms post-stimulus for qualitative assessment of developmentally changing template magnitudes across age groups. Colour bar indicates the template magnitude.

The template fit to the average EEG response is displayed for each stimulus modality at each of the three age categories (Figure 6B timeseries). In general, the visual and tactile templates fit the EEG data well for all age groups. However, the noxious template fit poorly for the youngest ages (PMA < 32 weeks). A subset of these infants exhibited the positive deflection that the noxious template is designed to fit, while another subset exhibited delta brush responses. Delta brush activity was defined as a slow delta wave with superimposed alpha or beta rhythms (André et al., 2010). These delta brush responses were only seen in response to noxious stimulation in some of the youngest infants (see triangle markers in Figure 6A Noxious). Due to the mix of positive deflection and delta brush responses, the noxious template magnitude at PMA < 32 weeks has a group average of approximately zero, suggesting the noxious-evoked delta brush activity is reducing at this developmental period and the positive deflection response is becoming dominant.

Fitting the templates to all electrodes across the scalp, the developmentally changing spatial patterns of the stimulus-evoked deflections were qualitatively assessed across age groups (Figure 6B heat maps). For visual and noxious evoked template magnitudes, which both positively correlated with PMA, the spatial distribution of evoked activity became increasingly localised around the dominant electrode (Oz for visual, Cz for noxious) as age and magnitude increased. Conversely, for tactile evoked template magnitudes, which were negatively correlated with PMA, the spatial distribution of evoked activity became decreasingly localised around the dominant Cz electrode as age increased and magnitude decreased.

### 3.3. The impact of premature extrauterine development on stimulus-evoked brain activity is detectable using template magnitudes

To assess the impact of premature extrauterine exposure on sensory development, we compared stimulus-evoked template magnitudes between a very preterm group and a late preterm group. These two groups were matched for PMA (gestational age + postnatal age), but the very preterm group had been developing postnatally for an average of 44 days, while the late preterm group had been developing postnatally for an average of 6 days. Compared to the late preterm infants, the very preterm infants had significantly larger visual-evoked response magnitudes (Cohen’s d=1.287, p-value=0.02) and significantly smaller tactile-evoked response magnitudes (d=–1.016, p-value=0.026) (Figure 7A). There was no significant difference between the groups for the noxious-evoked response magnitudes (d=0.042, p-value=0.93).

**Figure 7:**
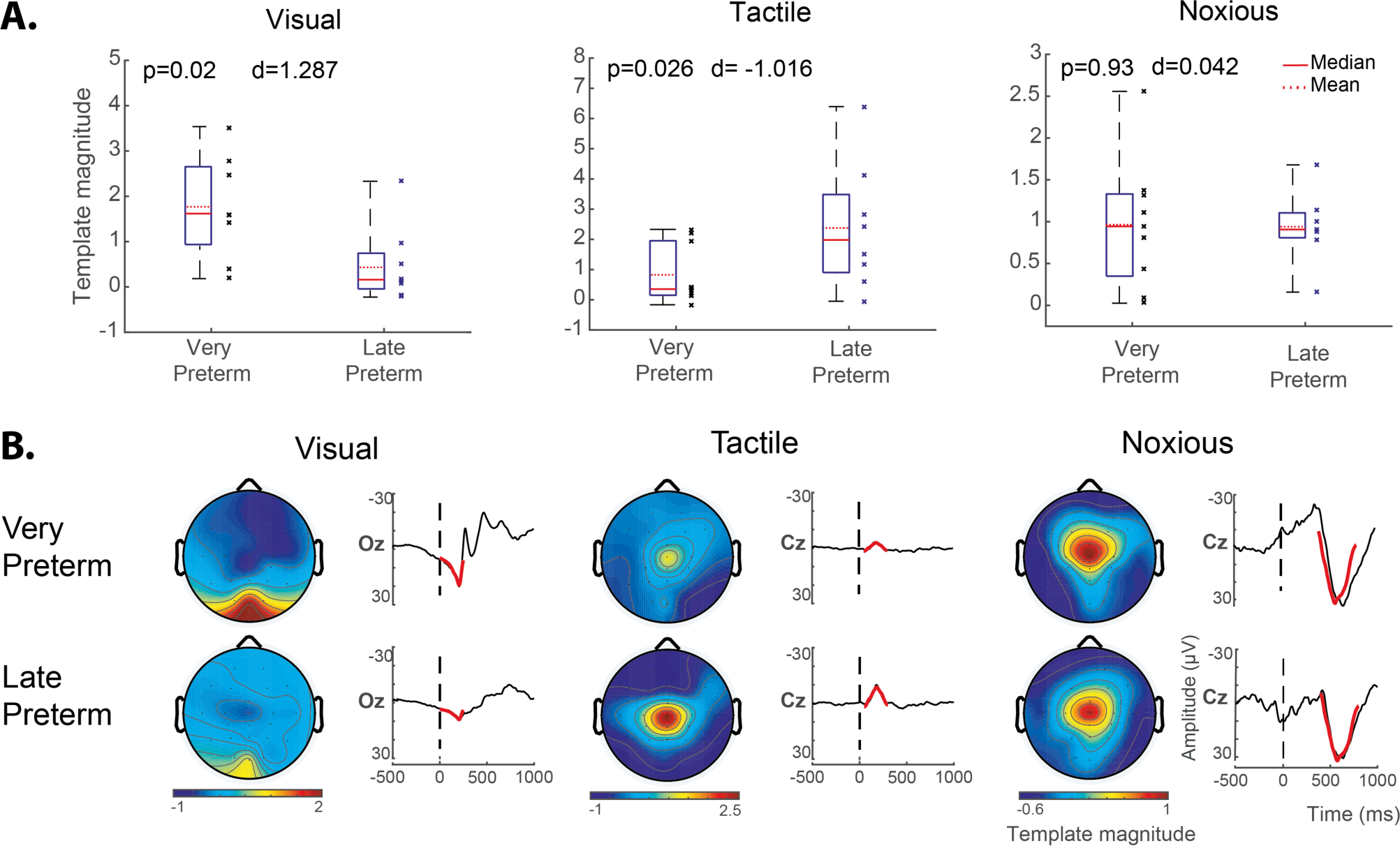
The impact of premature extrauterine development on stimulus-evoked response template magnitudes. Response magnitudes were assessed using the template analysis approach, and group average response magnitudes were compared between the very preterm infants (<32 weeks) and late preterm infants (34-36 weeks). Both groups were assessed at matched PMA of 34–37 weeks. Visual-evoked template magnitudes were calculated at electrode Oz, and tactile- and noxious-evoked template magnitudes were calculated at electrode Cz. (A) Quantitative comparison of evoked template magnitudes between groups using linear random effects regression model. Compared to the late preterm infants, the very preterm infants had significantly larger visual-evoked response magnitudes (left) and significantly smaller tactile-evoked response magnitudes (middle). There was no significant difference between the groups for the noxious-evoked response magnitudes (right). For all plots, d = Cohen’s d displaying the standardised difference in mean magnitudes; p = two-sided p-values. (B) Heat maps display the spatial patterns of the stimulus-evoked deflections, for qualitative assessment between groups. The timeseries display the templates (in red) superimposed onto the average EEG response (black) 500 ms pre-stimulus and 1000 ms post-stimulus for qualitative assessment of variable response magnitudes between groups. Colour bar indicates the template magnitude scaling. For all stimulus modalities, the templates fit the data well in both groups, and response topography patterns did not appear to be noticeably different between groups.

Given that each stimulus-evoked response template is scaled such that a template magnitude equal to one is the expected value for full term healthy infants (see methods section 2.2.5.), the average absolute value of the template magnitudes for the very preterm and late preterm infants in Figure 7A can be compared to this benchmark value of one. Thus relative to the average full term healthy cohort template magnitudes, the very preterm infants exhibited larger visual-evoked magnitudes (mean = 1.77) and smaller tactile-evoked magnitudes (mean = 0.80); and the late preterm infants exhibited smaller visual-evoked magnitudes (mean = 0.48) and larger tactile-evoked magnitudes (mean = 2.39).

The template fit to the average EEG response is displayed for each stimulus modality for both infant groups (Figure 7B timeseries). In general, the visual, tactile, and noxious templates fit the EEG data well for both groups. Fitting the templates to all electrodes across the scalp, differences in spatial patterns of the stimulus-evoked deflections were qualitatively assessed between groups (Figure 7B heat maps). The topography of response magnitudes did not appear to differ between groups.

## 4. Discussion

This study demonstrates that premature extrauterine exposure has a significant impact on the neural development of visual and tactile systems, detectable in peri-term aged infants and assessed using EEG-recorded brain activity in response to sensory stimulation. Features of the evoked brain activity were identified using a standardised template analysis approach that is a generalisation of a methodology initially developed for the assessment of noxious-evoked EEG brain activity (Hartley et al., 2017). The standardisation of the analysis minimised analytic heterogeneity across the stimulus modalities, and the selection of dominant features of the response signal addressed issues of low statistical power that often accompany infant studies of brain function development due to limited sample sizes and number of stimulus trials e.g. single-trial clinically required painful procedures.

A potential challenge to using the current data-driven approach to feature extraction, which relies on large signal amplitude and survival of statistical thresholding, is the integration of novel template analysis results to previously established features of evoked potentials. The template waveforms identified in the present study for visual-, tactile-, and noxious-evoked activity characterised prominent signal deflections in well-defined temporal windows (Figure 5). Assessing deflection polarity, latency to peak amplitude, and correlation between template magnitude and infant age, the template features bear close correspondence to previously characterised evoked potentials for the three modalities (Slater et al., 2010b; Taylor et al., 1987; Whitehead et al., 2019). The visual template bears close correspondence to the P200 visual-evoked potential, which has an approximate mean peak latency of 211 ms by term age, and has been shown to increase in amplitude with infant age (Taylor et al., 1987). The tactile template bears close correspondence to the N2 somatosensory-evoked potential, which has an approximate mean peak latency of 160 ms (Whitehead et al., 2019). While the tactile-evoked amplitude of N2 to foot stimulation was shown to have a non-significant association with age, the hand stimulation-evoked N2 amplitude has been shown to decrease with infant age (Whitehead et al., 2019). Finally, the noxious template bears close correspondence to the P560 noxious-evoked potential (Slater et al., 2010b), also known as the P3 potential (Jones et al., 2017), which has an approximate mean peak latency of 560 ms (Slater et al., 2010b), and has been shown to increase in amplitude with infant age (Hartley et al., 2016). This consistency in polarities, latencies, and age correlations between the current template features and established stimulus-evoked potentials suggests the template analysis approach has selected prominent features of evoked brain activity that can integrate into the existing literature and advance our understanding of the impact of premature exposure to the extrauterine environment on cortical sensory processing. The noxious template derived in the current study also bears a striking correspondence to our previously established noxious template (Hartley et al., 2017), highlighting its reproducibility and the robustness of the template analysis approach applied to the infant population.

Using this standardised template analysis approach, we observed that in longitudinally studied very preterm infants both the visual- and noxious-evoked template magnitudes increased, and tactile-evoked template magnitudes decreased with age, demonstrating the sensitivity of these evoked response features to developmental maturation (Figure 6). Further, in comparing age-matched infants born in the very preterm and late preterm periods, the very preterm infants had significantly larger visual-evoked and smaller tactile-evoked template magnitudes (Figure 7), demonstrating the impact of premature extrauterine exposure on both the visual and tactile systems. Given that each stimulus-evoked response template was normalised such that the expected value for full term healthy infants is one, it is interesting to note that the very prematurely born infants had visual responses on average greater than term infants and average tactile responses of lower magnitude than term-born infants. Taken together, these findings suggest that the infants with greater premature extrauterine exposure had accelerated neural functional maturation of both visual and tactile systems, assessed using EEG cortical evoked activity.

Accelerated or precocious maturation of neural function due to prematurity has previously been suggested for some sensory modalities as well as spontaneous resting-state activity. A longitudinal study has previously reported that preterm extra-uterine visual experience accelerates maturation of the VEP waveform (Tsuneishi and Casaer, 2000). By studying the decreasing latency of the stimulus-evoked activity with increasing maturation, extrauterine life has also been shown to accelerate maturation of the visual system (Schwindt et al., 2018; Taylor et al., 1987). Prematurity can also precipitate the early onset of binocular vision, assessed using visual evoked EEG activity (Jandó et al., 2012). Similarly, premature extrauterine life has been shown to accelerate maturation of the auditory system, assessed using latencies of evoked EEG activity (Cavalcanti et al., 2020). And using auditory verbal stimulation, haemodynamic responses assessed using NIRS had shorter latency as a consequence of early extrauterine exposure, suggesting accelerated maturation of the underlying neural responses, the subsequent haemodynamic response, the neurovascular coupling mechanism, or a combination thereof (Nishida et al., 2008). While these previous findings lend support to an accelerated maturation interpretation of the visual evoked activity observed in the current study, we were unable to assess the influence of prematurity on auditory-evoked cortical activity due to lack of statistically significant evoked responses (Figure 3). Future optimisation of the nature or delivery of the auditory stimulus will address this issue.

The attenuation of tactile-evoked response magnitudes found in infants born very prematurely in this study is consistent with previous infant studies. Although Maitre and colleagues (Maitre et al., 2017) studied a later potential (P2), they observed that ex-premature infants at term exhibit attenuated tactile-evoked EEG responses compared to term-born infants, which are proportional to the degree of prematurity at birth. A similar but non-significant trend in amplitude was also observed in another infant EEG study (Tombini et al., 2009) and a MEG study similarly reported significant reduction in tactile-evoked responses in ex-premature infants (Nevalainen et al., 2008). In all of these studies, it is unclear whether this represents precocious normal somatosensory development, which mirrors experience-dependent maturation that normally occurs postnatally in term-born infants, or whether these observed effects indicate alternative developmental trajectories. A future longitudinal study of multisensory evoked responses in term-born infants might allow us to address this question. Interestingly, in rats, the process of birth initiates the development of somatosensory map formation and this process is also initiated and accelerated by preterm birth (Toda et al., 2013).

There is little published regarding the influence of premature extrauterine exposure on noxious-evoked cortical activity in the peri-term period. To the best of the authors’ knowledge, only one EEG study has previously compared age-matched term and ex-premature infants (Slater et al., 2010a), and this study found that infants with greater premature extrauterine exposure exhibited larger noxious-evoked EEG responses. The magnitude of the noxious-evoked cortical activity has previously been shown to be sensitive to developmental maturation (Hartley et al., 2016), and this finding has been replicated here (Figure 6). These observations suggest the previous findings (Slater et al., 2010a) are consistent with the interpretation that premature extrauterine exposure may result in accelerated maturation of the noxious-evoked cortical response assessed using EEG. However, in the current study, we failed to replicate the observation of a significant difference in noxious-evoked template magnitude between the age-matched very preterm and late preterm infants (Figure 7).

This lack of a significant difference may be a false negative due to inadequate statistical power. Of the three stimulus modalities used in the cross-sectional study, only the noxious stimulus was a single-trial clinical heel lance, while the visual and tactile stimuli were trains of multi-trial experimental stimuli, likely resulting in the noxious-evoked activity being the noisiest most variable response. Alternatively, the lack of a significant difference may be a true negative – the effect of extra-uterine experience on noxious-evoked responses may be dependent on the degree of exposure to postnatal stressors and insults. In the current study, the ten prematurely-born infants studied in our very preterm cohort were remarkably healthy, with no significant intracranial pathology, no history of surgery, a low burden of pain exposure, and only two infants requiring invasive ventilation for >24 hours Conversely, the previous study included infants with significant neurological pathology (intraventricular haemorrhages grades 1-4) and greater exposure to invasive ventilation and painful clinical procedures including surgery. Future studies using the standardised template analysis approach directly contrasting two populations of prematurely-born infants matched for both gestational and postnatal ages should explore these various environmental and pathological factors that may impact the maturation of noxious-evoked brain activity.

The influence of premature extrauterine exposure on brain function is not limited to stimulus-evoked activity, but also impacts on resting-state activity. Using amplitude-integrated EEG assessment of ongoing spontaneous brain activity to compare age-matched infants with varying gestational ages, infants born more prematurely displayed advanced maturation compared to less premature infants (Soubasi et al., 2009). Ex-premature infants at term also display some features of accelerated maturation of EEG-sleep organisation, having longer sleep cycles, less fragmentation, and more quiet sleep than their term-born counterparts (Scher, 1997), which may be an adaptive response to the environment but representative of dysmaturity. Resting-state fMRI has also been used to assess the impact of premature extrauterine life and differences in functional connectivity are observable between extrauterine premature infants and in-utero foetuses (De Asis-Cruz et al., 2020). Specifically, the extrauterine infants had greater functional connectivity between brain regions within the auditory (temporal regions), visual (occipital regions), and stress/pain (hippocampal and insular regions) systems, suggesting accelerated maturation of these systems due to extrauterine exposure.

There are several conceptual models in the literature proposed to explain accelerated or precocious maturation of neural function due to premature extrauterine exposure. The stress acceleration hypothesis (Callaghan and Tottenham, 2016), predictive adaptive and other adaptive response hypotheses (Gluckman et al., 2005; Sih, 2011) propose that early life stressors such as premature exposure to the extrauterine environment could result in accelerated maturation along normal developmental trajectories or the adoption of alternative developmental trajectories to adapt to the challenging extrauterine context. They further suggest that the short-term benefits of developmental acceleration and adaptation may not hold long-term and are likely to incur numerous costs. The long-term neurodevelopmental deficits resulting from very preterm birth are well documented and range from motor deficits and cerebral palsy to cognitive impairments and social and emotional difficulties (Anderson and Doyle, 2004; Bhutta et al., 2002; Holsti et al., 2002; Laptook et al., 2005; Marlow et al., 2005; Nosarti et al., 2012; Saigal and Doyle, 2008; Taylor et al., 2004). The short-term costs in neural function detectable using fMRI at term-equivalent age for infants born prematurely have also been identified as widespread developmental delays and impairments in functional connectivity and global network measures of integration, segregation, and modularity (Bouyssi-Kobar et al., 2019; Eyre et al., 2021; Scheinost et al., 2016).

The complex variability in reported influences of premature exposure on infant brain function are likely due to heterogeneity across functional system development, brain activity measurement modality, and analysis methodology. Maturation of the infant brain’s neural systems is both spatially and functionally heterogeneous in timing and rate of development (Cao et al., 2017; Dubois et al., 2014, 2008; Ouyang et al., 2019) resulting in variability in system critical periods and consequent vulnerabilities (Reh et al., 2020). Additionally, unlike EEG and MEG, haemodynamic-based brain assessment modalities such as fMRI and NIRS will not only reflect the influence of prematurity on neural activity but also the impact on cerebral vascular and neurovascular coupling development (Kozberg and Hillman, 2016). Methodological heterogeneity across studies further compounds the challenges in pooling these observations into a unified framework of understanding – see (Botvinik-Nezer et al., 2020) for an informative example of the substantial effects that analytical flexibility can have on scientific conclusions. The approach taken in the current study aimed to address some of these limitations by studying multiple sensory modalities within the same infants, assessing evoked response features both longitudinally and cross-sectionally, while using a standardised analysis methodology across sub-studies. However, interpretational limitations still remain, which future multi-imaging modality studies may address. While simultaneous EEG and fMRI recordings in infants is incredibly challenging, it is possible (Arichi et al., 2017) and would be highly valuable to better understand the associations between prematurity-related effects on infant neurodynamics assessed using EEG and haemodynamics assessed using fMRI, and follow-up childhood sensorimotor and cognitive outcomes to assess the long-lasting effects of these peri-term age brain function differences, if any.

The identification of brain activity features sensitive to the impact of premature extrauterine exposure on the visual and tactile systems highlights the importance of brain-based measures in assessing healthy and disrupted neurodevelopment. The use of EEG features as outcome measures for assessing the efficacy of developmental care interventions is increasing in uptake (Burke, 2018; Neel et al., 2019). Findings consistently indicate positive differences in EEG activity in favour of the intervention group (Als et al., 2011; Kaffashi et al., 2013; McAnulty et al., 2013, 2009; Myers et al., 2015; Welch et al., 2014), and this consistency in findings suggests that neurodevelopmental differences may be detectable neurophysiologically before they manifest behaviourally (Burke, 2018). Previous studies have primarily focused on EEG features of spontaneous brain activity, such as coherence (Als et al., 2011; McAnulty et al., 2013, 2009; Myers et al., 2015), entropy (Kaffashi et al., 2013), and power (Welch et al., 2014). The current study outlines a standardised analysis methodology and outcome measures that can be used for multi-modal EEG evoked activity analysis for a more direct and targeted assessment of multiple sensory system function within an individual infant. Given the sensitivity of the premature infant’s neurodevelopment to sensory input (Colonnese et al., 2010; Milh et al., 2007; Wess et al., 2017), and the negative impact prematurity can have across sensory systems of the brain (Wallois et al., 2020), development and application of multi-modal EEG sensory assessment, such as the template analysis of evoked activity, is likely to be a valuable approach to complement resting-state EEG and behavioural assessments when studying the influence of prematurity on neurodevelopment and the efficacy of developmental care interventions.

## 5. Conclusions

We outline a standardised template analysis approach to measure the EEG dominant waveform magnitude evoked by visual, tactile, and noxious stimulation. We observe that the evoked template magnitudes for these three sensory modalities are sensitive to the infant’s age at the time of assessment, and that significant differences in neural processing of both visual and tactile stimulation exist between infants born very preterm and late preterm, demonstrating the impact of premature exposure to the extrauterine environment on brain function. Taken together, our findings suggest that premature extrauterine exposure may accelerate maturation of visual and tactile neural systems, and highlight the importance of brain-based EEG measures in assessing healthy and disrupted neurodevelopment. Additionally, we suggest that multi-modal evoked activity analysis, such as the template analysis approach, could complement resting-state EEG and behavioural assessments of the efficacy of developmental care interventions by providing direct and targeted assessment of multiple sensory systems within an individual infant.

## Data and code availability statement

The data that support the study findings are available from the corresponding author upon reasonable request. Due to ethical restrictions, it is appropriate to monitor access and usage of the data as it includes highly sensitive information. Data sharing requests should be directed to. The code underpinning template construction is available on GitLab: https://gitlab.com/paediatric_neuroimaging/stim-template-development. Template fitting to novel data and linear modelling, as detailed in methods, was performed using standard MATLAB commands (https://mathworks.com/products/matlab.html).

## CRediT authorship contribution statement

**Gabriela Schmidt Mellado**: Conceptualization, Methodology, Investigation, Data curation, Writing – Original draft, Visualization. **Kirubin Pillay**: Methodology, Software, Formal analysis, Data curation, Writing – Original draft, Visualisation. **Eleri Adams**: Writing – Review & Editing, Supervision. **Ana Alarcon**: Conceptualization, Writing – Review & Editing, Supervision. **Foteini Andritsou**: Investigation, Writing – Review & Editing. **Maria M Cobo:** Investigation, Writing – Review & editing. **Ria Evans Fry**: Investigation, Writing – Review & Editing. **Sean Fitzgibbon:** Writing-Review &editing. **Fiona Moultrie**: Writing – Review & Editing. **Luke Baxter**: Project administration, Validation, Writing – Original draft. **Rebeccah Slater**: Conceptualization, Writing – Review & Editing, Supervision, Project administration, Funding Acquisition.

## Data Availability

The data that support the study findings are available from the corresponding author upon reasonable request. Due to ethical restrictions, it is appropriate to monitor access and usage of the data as it includes highly sensitive information. Data sharing requests should be directed to.

## Acknowledgements

We thank the neonates and their parents for generously taking part in this study. We thank Marianne van der Vaart, Miranda Buckle, Deniz Gursul, and Shellie Robinson for their assistance during data collection. This study was funded by a Senior Wellcome Research Fellowship awarded to R.S. (207457/Z/17/Z).

